# Exposure to autoimmune disorders increases Alzheimer’s disease risk in a multi-site electronic health record analysis

**DOI:** 10.1101/2024.05.02.24306649

**Authors:** Grace D. Ramey, Alice Tang, Thanaphong Phongpreecha, Monica M. Yang, Sarah R. Woldemariam, Tomiko T. Oskotsky, Thomas J. Montine, Isabel Allen, Zachary A. Miller, Nima Aghaeepour, John A. Capra, Marina Sirota

**Author notes:** Corresponding authors information: John A. Capra, UCSF Bakar Computational Health Sciences Institute, San Francisco, CA 94143; Marina Sirota, UCSF Bakar Computational Health Sciences Institute, San Francisco, CA 94143.

## Abstract

Molecular studies of Alzheimer’s disease (AD) implicate potential links between autoimmunity and AD, but the underlying clinical relationships between these conditions remain poorly understood. Electronic health records (EHRs) provide an opportunity to determine the clinical risk relationship between autoimmune disorders and AD and understand whether specific disorders and disorder subtypes affect AD risk at the phenotypic level in human populations. We evaluated relationships between 26 autoimmune disorders and AD across retrospective observational case-control and cohort study designs in the EHR systems at UCSF and Stanford. We quantified overall and sex-specific AD risk effects that these autoimmune disorders confer. We identified significantly increased AD risk in autoimmune disorder patients in both study designs at UCSF and at Stanford. This pattern was driven by specific autoimmunity subtypes including endocrine, gastrointestinal, dermatologic, and musculoskeletal disorders. We also observed increased AD risk from autoimmunity in both women and men, but women with autoimmune disorders continued to have a higher AD prevalence than men, indicating persistent sex-specificity. This study identifies autoimmune disorders as strong risk factors for AD that validate across several study designs and EHR databases. It sets the foundation for exploring how underlying autoimmune mechanisms increase AD risk and contribute to AD pathogenesis.

## Introduction

Alzheimer’s disease (AD) is a debilitating neurodegenerative disease that is accompanied by enormous social and economic burdens, and its prevalence is increasing due to the growing aging population worldwide (1,2). AD is characterized biologically by amyloid plaques and tau deposition in the brain, while clinical syndromic diagnoses, such as specific forms of progressive memory loss, have evolved with the development of better AD diagnostic tests and biomarkers (3,4). Treatments that slow cognitive decline have been a large focus of AD research over the past several years (5), and understanding of underlying risks and pathogenesis in order to treat the disease at an earlier stage is especially important considering that current treatments are still unable to fully rescue normal cognition (6).

Many prior molecular and genetic studies suggest a potential role of the immune system and chronic inflammation in AD pathogenesis (7–9). Indeed, over half of the genetic variants associated with AD to date are primarily expressed in immune cells (10). Furthermore, several studies point to immune pathways like the NLRP3 inflammasome (11) and complement system (12–14) becoming dysregulated in AD experimental animal and human models. However, the extent of contribution of immune system dysfunction to AD remains poorly understood at the clinical phenotype level in diverse human populations. Autoimmune disorders are one potential source of chronic immune dysregulation, and their clinical risk relationship with neurodegenerative diseases like AD has yet to be fully characterized. Furthermore, autoimmune disorders exhibit a similar sex disparity to AD (15,16), affecting women more so than men (17–19), suggesting a potential relationship between biological mechanisms and clinical manifestations that has yet to be quantified. Therefore, studying risk relationships in individuals with autoimmune disorders and AD will provide a powerful way to understand the role of autoimmunity as a risk factor for AD overall and across sexes.

With advances in curation of real-world datasets (20), such as electronic health records (EHRs), there is a great opportunity to investigate clinical risk relationships between many autoimmune disorders and AD. The large sample sizes that EHRs provide also allow for robust analyses that can be stratified in a sex- and disease-specific manner and validated across hospital sites. Here, we examine risk associations between AD and 26 different autoimmune disorders in the UCSF EHR system, and we further stratify our analyses by sex to holistically understand the biological effects of immune dysfunction on AD pathogenesis at the phenotypic level. We show that there is a clear and strong risk association between autoimmune disorders and AD overall and in both men and women, but that women with autoimmune disorders continue to have the highest AD prevalence compared to men with autoimmune disorders. We show evidence for increased AD risk effects from particular autoimmune disorders and disorder subtypes, and we further investigate the timing of AD onset in autoimmune disorders patients. Finally, we provide robust validation of our risk associations from the Stanford EHR system to demonstrate stability of risk signals across different study designs and mitigate potential confounding factors.

## Results

We selected patients with autoimmune disorders and/or AD for case-control and cohort study designs from the UCSF and Stanford EHR databases, which contain information on over 5 million and 3.8 million patients, respectively. For patients with autoimmune disorders, we identified individuals with each of 26 different autoimmune disorders of interest (Table S1), and type 1 diabetes, rheumatoid arthritis, autoimmune thyroiditis, and inflammatory bowel disease were among the most prevalent in the study groups. The case-control study groups consisted of 7,812 individuals (3,906 AD patients and 3,906 non-AD controls, Fig 1, S1) from UCSF and 13,292 individuals (6,646 AD patients and 6,646 non-AD controls, Fig 1, S1) from Stanford. The cohort study groups consisted of 27,630 individuals (13,815 autoimmune disorder patients and 13,815 non-autoimmune controls, Fig 1, S1) from UCSF and 260,516 individuals (130,258 autoimmune disorder patients and 130,258 non-autoimmune controls, Fig 1, S1) from Stanford. In the UCSF data set, the mean (±SD) lifespan in the case-control group was 80.05 (±6.74) years for AD patients and 80.07 (±6.73) years in non-AD controls, while in the cohort study group, the mean lifespan for autoimmunity patients was 69.25 (±13.21) years and 69.10 (±13.38) years for non-autoimmune controls. Because individuals had a higher proportion of censored death information in the Stanford data set, these people were assessed by birth year rather than lifespan, and we verified that this difference would not significantly influence results (see Methods and Sensitivity Analyses). In the Stanford data set, the mean birth year (±SD) was 1934.53 (±10.11) for AD patients and 1934.50 (±10.07) for non-AD controls in the case-control study group, while in the cohort study group, the mean birth year both for autoimmunity patients and non-autoimmune controls was 1968.38 (±21.83) (Table 1). Women made up a majority of each of our study groups, representing 61.9% (case-control) and 57.9% (cohort) of individuals in the UCSF study groups, and 62.3% (case-control) and 64.9% (cohort) of individuals in the Stanford study groups. Further demographic information across study designs and EHR data sets is listed in Table 1.

**Figure 1.**
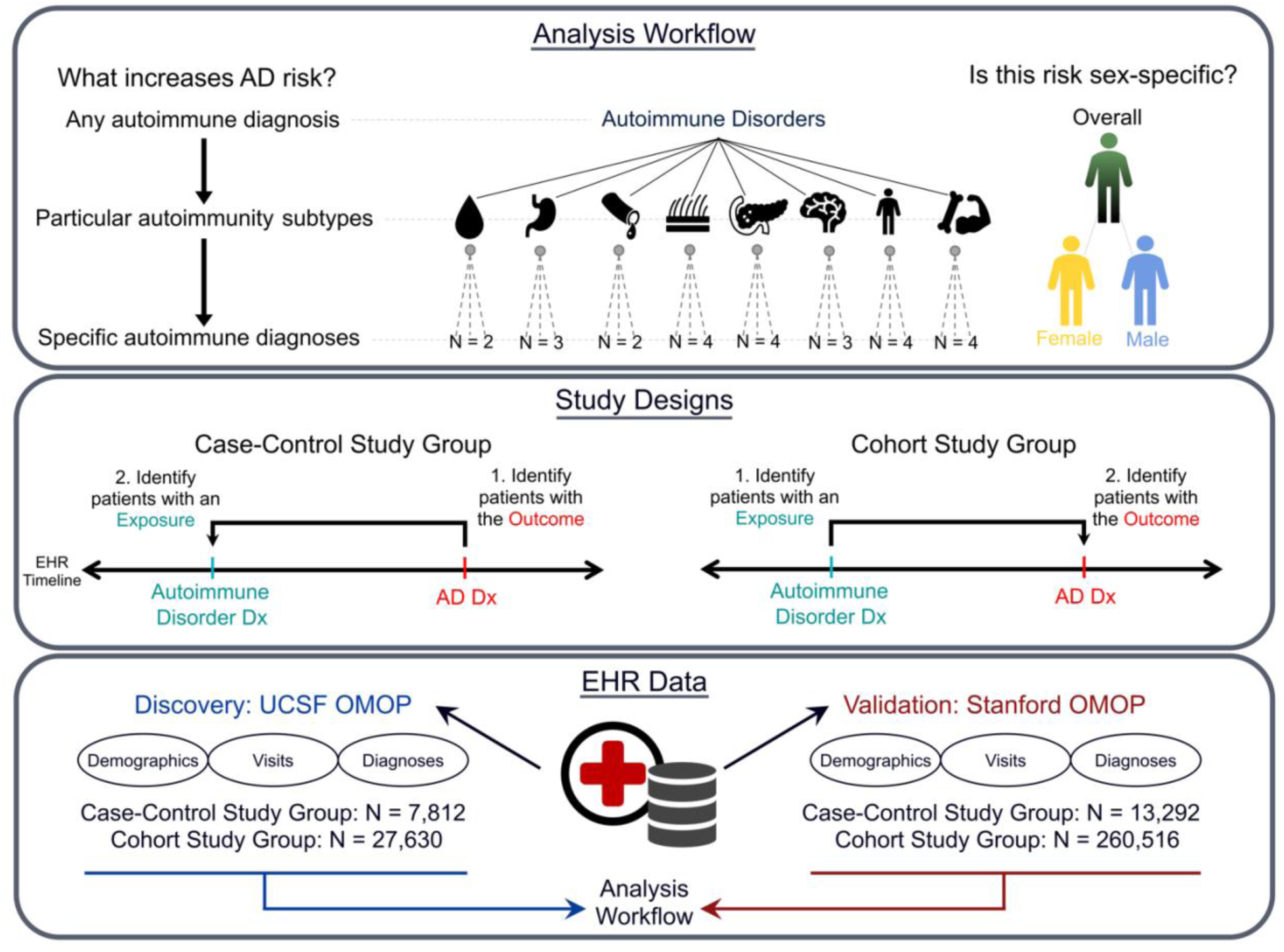
Risk analysis framework and observational study designs. We take a top-down hierarchical approach to calculate AD risk in autoimmune disorder patients based on two large electronic health record (EHR) data sets. We compute odds ratios to assess if any autoimmune disorder diagnosis, a particular autoimmune subtype diagnosis, or a specific autoimmune disorder diagnosis increases AD risk. We used both a case-control and cohort study design to ensure robustness and reduce biases. For the case-control study design, we first identified patients with the *outcome* of interest (an AD diagnosis, red) and then determined which of the AD patients also had an autoimmune diagnosis (blue). We matched the AD patients to non-AD controls using propensity score matching and gathered information on other demographic variables for cases and controls. For the cohort study design, we identified patients with the *exposure* first (an autoimmunity diagnosis, blue) and determined which of the exposed patients also had an AD diagnosis (red). Demographic information was extracted and propensity score matching was used to match autoimmune cases to non-autoimmune controls. We used these study structures to analyze data from both the UCSF (discovery) and Stanford (validation) EHR databases. Dx = diagnosis

**Table 1.** UCSF and Stanford Study Group Demographics.

| UCSF |  |  |  |  | Stanford |  |  |  |  |
| --- | --- | --- | --- | --- | --- | --- | --- | --- | --- |
| Case-Control Study Design |  |  |  |  | Case-Control Study Design |  |  |  |  |
|  | Non-AD Controls | AD Patients | p value | SMD |  | Non-AD Controls | AD Patients | p value | SMD |
| N | 3,906 | 3,906 |  |  | N | 6,646 | 6,646 |  |  |
| Sex (%) |  |  | 1 | <0.001 | Sex (%) |  |  | 1 | <0.001 |
| Female | 2416 (61.9) | 2416 (61.9) |  |  | Female | 4,143 (62.3) | 4,143 (62.3) |  |  |
| Male | 1,490 (38.1) | 1,490 (38.1) |  |  | Male | 2,503 (37.7) | 2,503 (37.7) |  |  |
| Unknown | 0 (0.0) | 0 (0.0) |  |  | Unknown | 0 (0.0) | 0 (0.0) |  |  |
| Self-reported race (%) |  |  | 1 | <0.001 | Self-reported race (%) |  |  | 1 | <0.001 |
| Asian | 531 (13.6) | 531 (13.6) |  |  | Asian | 952 (14.3) | 952 (14.3) |  |  |
| Black or African American | 232 (5.9) | 232 (5.9) |  |  | Black or African American | 315 (4.7) | 315 (4.7) |  |  |
| Native American or Alaska Native | 4 (0.1) | 4 (0.1) |  |  | Native American or Alaska Native | 9 (0.1) | 9 (0.1) |  |  |
| Native Hawaiian or Other Pacific Islander | 174 (4.5) | 174 (4.5) |  |  | Native Hawaiian or Other Pacific Islander | 39 (0.6) | 39 (0.6) |  |  |
| White | 2,634 (67.4) | 2,634 (67.4) |  |  | White | 4,182 (62.9) | 4,182 (62.9) |  |  |
| Other | 331 (8.5) | 331 (8.5) |  |  | Other | 813 (12.2) | 813 (12.2) |  |  |
| Unknown/Decline to State | 0 (0.0) | 0 (0.0) |  |  | Unknown/Decline to State | 336 (5.1) | 336 (5.1) |  |  |
| Self-reported ethnicity (%) |  |  | 1 | <0.001 | Self-reported ethnicity (%) |  |  | 1 | <0.001 |
| Not Hispanic or Latino | 3,672 (94.0) | 3,672 (94.0) |  |  | Not Hispanic or Latino | 5,732 (86.2) | 5,732 (86.2) |  |  |
| Hispanic or Latino | 234 (6.0) | 234 (6.0) |  |  | Hispanic or Latino | 503 (7.6) | 503 (7.6) |  |  |
| Unknown/Decline to State | 0 (0.0) | 0 (0.0) |  |  | Unknown/Decline to State | 411 (6.2) | 411 (6.2) |  |  |
| Birth year (mean (SD)) | 1935.52 (5.93) | 1935.52 (5.92) | 1 | <0.001 | Birth year (mean (SD)) | 1934.53 (10.11) | 1934.50 (10.07) | 0.827 | 0.004 |
| Lifespan, years (mean (SD)) | 80.05 (6.74) | 80.07 (6.73) | 0.945 | 0.002 | Lifespan, years (mean(SD)) | NA | NA |  |  |
| Autoimmune disease status = True (%) | 235 (6.0) | 377 (9.7) | <0.001 | 0.136 | Autoimmune disease status = True (%) | 415 (6.2) | 575 (8.7) | <0.001 | 0.092 |
| Cohort Study Design |  |  |  |  | Cohort Study Design |  |  |  |  |
|  | Non-Autoimmune Controls | Autoimmune Disorder Patients | p value | SMD |  | Non-Autoimmune Controls | Autoimmune Disorder Patients | p value | SMD |
| N | 13,815 | 13,815 |  |  | N | 130,258 | 130,258 |  |  |
| Sex (%) |  |  | 1 | <0.001 | Sex = Male (%) |  |  | 1 | <0.001 |
| Female | 7,994 (57.9) | 7,994 (57.9) |  |  | Female | 84,515 (64.9) | 84,515 (64.9) |  |  |
| Male | 5,821 (42.1) | 5,821 (42.1) |  |  | Male | 45,717 (35.1) | 45,717 (35.1) |  |  |
| Unknown/Decline to State | 0 (0.0) | 0 (0.0) |  |  | Unknown/Decline to State | 26 (0.0) | 26 (0.0) |  |  |
| Self-reported race (%) |  |  | 1 | <0.001 | Self-reported race (%) |  |  | 1 | <0.001 |
| Asian | 1,241 (9.0) | 1,241 (9.0) |  |  | Asian | 19,022 (14.6) | 19,022 (14.6) |  |  |
| Black or African American | 1,195 (8.7) | 1,195 (8.7) |  |  | Black or African American | 5,036 (3.9) | 5,036 (3.9) |  |  |
| Native American or Alaska Native | 85 (0.6) | 85 (0.6) |  |  | Native American or Alaska Native | 559 (0.4) | 559 (0.4) |  |  |
| Native Hawaiian or Other Pacific Islander | 541 (3.9) | 541 (3.9) |  |  | Native Hawaiian or Other Pacific Islander | 1,087 (0.8) | 1,087 (0.8) |  |  |
| White | 8,916 (64.5) | 8,916 (64.5) |  |  | White | 72,757 (55.9) | 72,757 (55.9) |  |  |
| Other | 1,837 (13.3) | 1,837 (13.3) |  |  | Other | 22,111 (17.0) | 22,111 (17.0) |  |  |
| Unknown/Decline to State | 0 (0.0) | 0 (0.0) |  |  | Unknown/Decline to State | 9,686 (7.4) | 9,686 (7.4) |  |  |
| Self-reported ethnicity (%) |  |  | 1 | <0.001 | Self-reported ethnicity (%) |  |  | 1 | <0.001 |
| Not Hispanic or Latino | 12,339 (89.3) | 12,339 (89.3) |  |  | Not Hispanic or Latino | 101,442 (77.9) | 101,442 (77.9) |  |  |
| Hispanic or Latino | 1,476 (10.7) | 1,476 (10.7) |  |  | Hispanic or Latino | 17,816 (13.7) | 17,816 (13.7) |  |  |
| Unknown/Decline to State | 0 (0.0) | 0 (0.0) |  |  | Unknown/Decline to State | 11,000 (8.4) | 11,000 (8.4) |  |  |
| Birth year (mean (SD)) | 1946.49 (13.43) | 1946.55 (13.51) | 0.705 | 0.005 | Birth year (mean (SD)) | 1968.38 (21.83) | 1968.38 (21.83) | 0.724 | 0.001 |
| Lifespan, years (mean (SD)) | 69.25 (13.21) | 69.10 (13.38) | 0.327 | 0.012 | Lifespan, years (mean(SD)) | NA | NA |  |  |
| AD status = True (%) | 195 (1.4) | 379 (2.7) | <0.001 | 0.093 | AD status = True (%) | 354 | 579 | <0.001 | 0.029 |

Reports of 'NA' for lifespan indicate censored death information in the Stanford study groups. SMD = Standardized Mean Difference, SD = Standard deviation

We compared the risk of being diagnosed with AD in patients with autoimmune disorders compared to non-autoimmune controls across case-control and cohort study designs in both the discovery (UCSF) and validation (Stanford) data sets. We evaluated the risk of AD in the study groups both overall and in a sex-stratified manner across multiple levels of autoimmune disorder stratifications to determine specific autoimmune drivers of AD risk.

### Autoimmune disorder diagnoses are significantly associated with increased risk of AD overall and within sex-specific groups

In the case-control study groups, overall, individuals with autoimmune disorders had significantly higher odds of an AD diagnosis compared to non-autoimmune controls in both discovery (OR = 1.7, 95% CI 1.4-2.0, p = 2.5e-9, Fig 2A) and validation (OR = 1.4, 95% CI 1.2-1.6, p = 1.4e-7, Fig 2A) data sets. We observed even larger AD risk associations with autoimmune disorders in the cohort study groups, where odds ratios were 2.0 (95% CI 1.7-2.4, p = 7.0e-15, Fig 2A) and 1.6 (95% CI 1.4-1.9, p = 1.6e-13, Fig 2A) for UCSF and Stanford, respectively. This consistent elevated risk across study designs and EHR systems highlights a strong connection between autoimmunity and AD.

**Figure 2.**
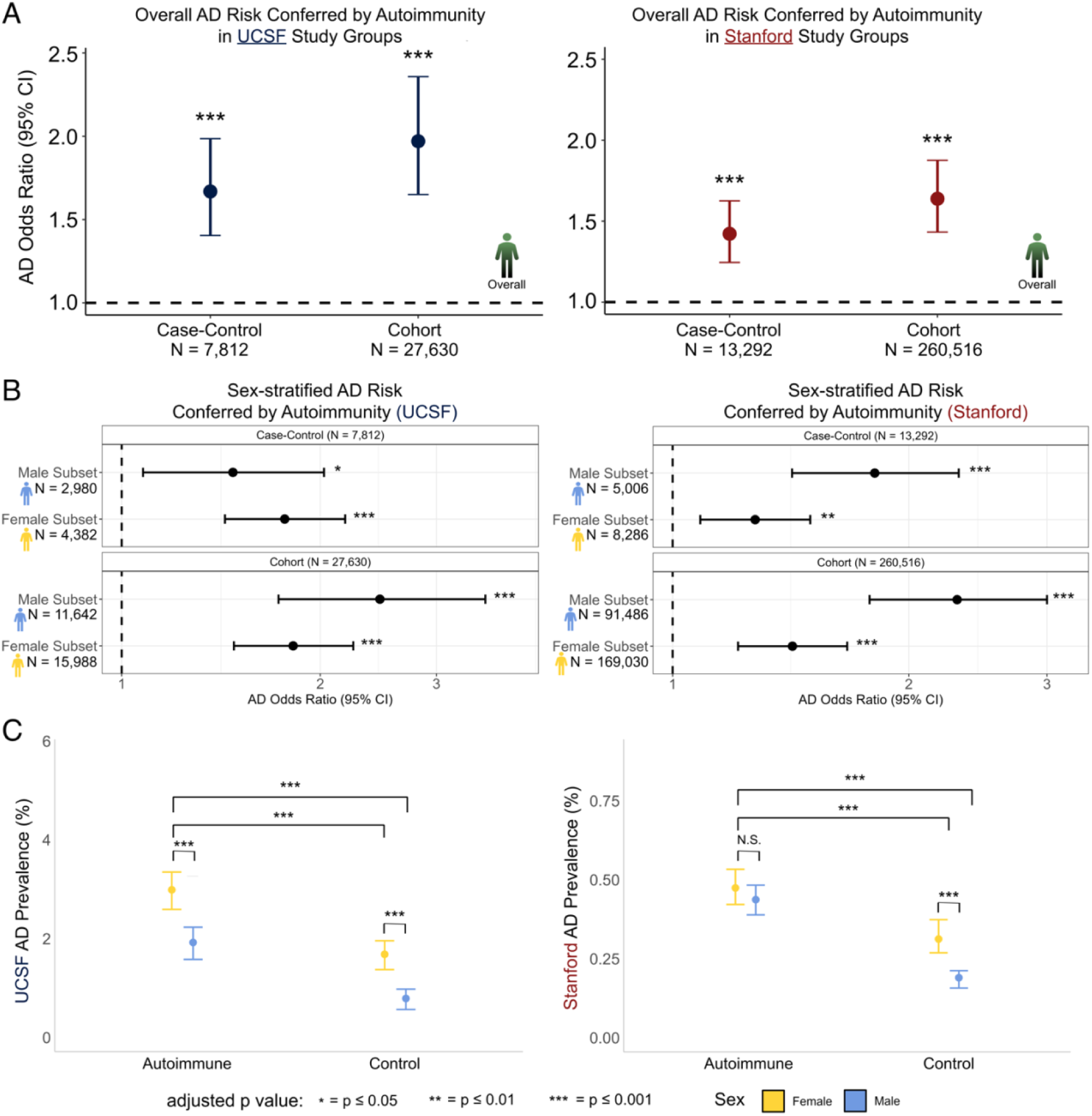
Autoimmune disorders are associated with increased AD risk across study designs and EHR data sets. **A)** Odds ratios quantifying AD risk in autoimmune patients versus non-autoimmune controls. We observed increased odds of AD across both the UCSF (left) and Stanford (right) data sets, and across case-control and cohort study groups within each data set, robustly highlighting greater AD risk conferred by autoimmunity. **B)** AD risk in the female- and male-only subsets of each data set. Increased AD risk was present in both sexes in each data set. **C)** AD prevalence calculated in the cohort study designs in different sex and disorder strata. Confidence intervals were obtained by bootstrapping the data. AD prevalence was higher in women with autoimmune disorders compared to all other groups in both the UCSF (left) and Stanford (right) data.

We divided our discovery and validation data into female- and male-only subsets to determine if AD risk remained elevated within sex-specific groups and learn if the overall risk association was primarily driven by women. In the female-only subsets of the case-control study designs, we observed significantly greater odds of an AD diagnosis in women with autoimmune disorders compared to control women at both UCSF (OR = 1.8, 95% CI 1.4-2.2, p = 4.1e-8, Fig 2B) and Stanford (OR = 1.3, 95% CI 1.1-1.5, p = 3.0e-3, Fig 2B). Similarly, among women in the cohort study groups, we observed significantly greater AD risk in women with autoimmune disorders at UCSF (OR = 1.8, 95% CI 1.5-2.2, p = 4.9e–9, Fig 2B) and at Stanford (OR = 1.4, 95% CI 1.2-1.7, p = 1.2e-5, Fig 2B). There were also strong associations between autoimmunity and AD in the male-only subsets. In the case-control study groups, men with autoimmune disorders had significantly greater AD risk compared to control men across UCSF and Stanford studies (UCSF OR = 1.5, 95% CI 1.1-2.0, p = 0.014; Stanford OR = 1.8, 95% CI 1.4-2.3, p = 8.2e-7; Fig 2B). The results in the cohort study groups agreed (UCSF OR = 2.5, 95% CI 1.7-3.6, p=1.3e-7; Stanford OR = 2.3, 95% CI 1.8-3.0, p = 2.0e-11; Fig 2B), highlighting higher risk in men with autoimmune disorders compared to control men across both EHR systems. The increased AD risk observed in both men and women with autoimmune disorders suggests that increased AD risk conferred by autoimmunity is not driven solely by one sex.

### AD prevalence is increased in the presence of autoimmune disorders, but women remain the most affected

While autoimmunity associates with AD risk within both sexes, we next tested whether it elevated risk more in one sex than the other. Furthermore, we wanted to determine if the presence of autoimmunity diminishes the well-documented AD sex-disparity wherein women tend to develop AD more often than men. To address these questions, we conducted an AD prevalence analysis within the cohort study groups of our discovery and validation data sets. Due to the smaller number of men compared to women in our data, we conducted 1:1 matching of women to men based on demographic variables (see Methods) and computed AD prevalence across sex and autoimmunity stratifications. In our discovery data set, women with autoimmune disorders (N = 5,821) had the highest AD prevalence at 3.0%, followed by men with autoimmune disorders (N = 5,821) at 1.9%, control women (N = 5,821) at 1.7%, and finally control men (N = 5,821) at 0.79% (Fig 2C). While the absolute prevalence values were lower at Stanford, likely due to younger patients being included in the cohort study group as a result of censored age information (see Methods), there was a similar hierarchy in the validation data set, where women with autoimmune disorders (N = 45,743) had the highest AD prevalence at 0.47%, followed by men with autoimmune disorders (N = 45,743) at 0.43%, control women (N = 45,743) at 0.31%, and control men (45,743) at 0.19%. As expected, the higher prevalence in control women compared to control men (UCSF corrected p = 6.2e-5; Stanford corrected p = 1.1e-3; Fig 2C) corroborates well-documented sex-disparities in AD. We then discovered that women with autoimmune disorders exhibited a higher AD prevalence than men with autoimmune disorders in the UCSF data set (corrected p = 9.9e-4). The AD prevalence difference between autoimmunity patients of different sexes was roughly equal in magnitude to the difference between control patients of different sexes (1.1% versus 0.91%), suggesting that sex-disparities in AD remain present, even with autoimmunity conferring greater risk in both sexes. The intersex comparison in autoimmune patients in the Stanford data set was not significant (p = 0.4), but women with autoimmune disorders did exhibit a slightly higher AD prevalence than the corresponding men, likely indicating that women continue to bear the most risk for AD even when a significant immune perturbation like autoimmunity is at play.

### Specific autoimmune disorder subtypes are associated with greater AD risk, driven by individual disorders

Next, to determine if particular classes of autoimmune disorders are associated with AD risk more than others, we divided the 26 autoimmune disorders into subtypes based on the organ system each one primarily affects. This resulted in eight disease subtype categories: musculoskeletal, gastrointestinal, dermatologic, systemic, vascular, hematologic, neurologic, and endocrine (Fig 3A). We computed the AD odds ratios for individuals with these disease subtypes in each of the case-control and cohort study designs across UCSF and Stanford data sets. In the overall case-control and cohort study designs of the UCSF data set, autoimmune disorders in the gastrointestinal (case-control OR = 6.1, 95% CI 2.7-14.6, corrected p=7.2e-6; cohort OR = 2.5, 95% CI 1.4-4.7, corrected p=6.5e-3, Fig 3B), hematologic (case-control OR = 18.1, 95% CI 4.7-84.8, corrected p=2.3e-6; cohort OR = 4.5, 95% CI 1.8-13.4, corrected p=3.6e-3, Fig 3B), endocrine (case-control OR = 2.8, 95% CI 1.8-4.4, corrected p=6.0e-6; cohort OR = 1.9, 95% CI 1.4-2.6, corrected p =2.2e-4, Fig 3B), musculoskeletal (case-control OR = 2.5, 95% CI 1.7-3.9, corrected p=2.7e-5; cohort OR = 2.0, 95% CI 1.5-2.8, corrected p=4.7e-5), and dermatologic (case-control OR = 3.1, 95% CI 1.6-6.1, corrected p=1.2e-3; cohort OR = 2.4, 95% CI 1.4-4.0, corrected p=2.6e-3) categories associated with increased risk for AD. Systemic, vascular, and neurologic disease subtypes did not significantly associate with increased AD, potentially because we were underpowered to detect risk associations for these particular subtypes (Table S2).

**Figure 3.**
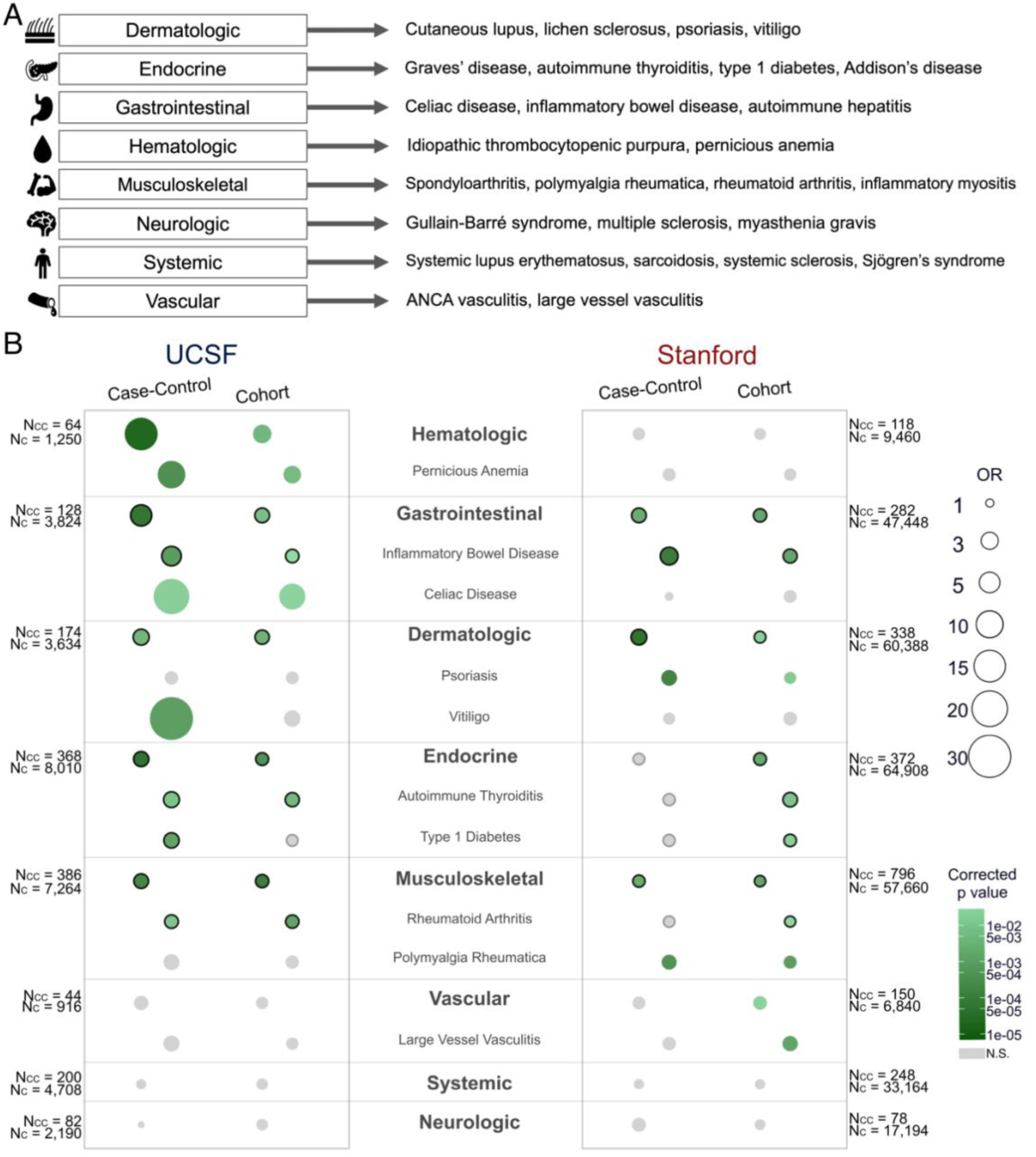
Specific autoimmune disorder subtypes and individual diseases associate with increased AD risk. **A)** Individual autoimmune disorders can be grouped into subtypes based on physiological symptomatology. We used eight subtype groups in our analysis. **B)** Odds ratios quantifying AD risk in patients with different autoimmune disorder subtypes and specific autoimmune disorders compared to controls. Disorder subtypes are in bold, and the specific disorders that fall into each subtype category are listed below. Only individual autoimmune disorders that were statistically significant are pictured. The larger and darker the circle is, the greater the effect size and significance of the odds ratio, respectively. N.S. = Not Significant after multiple testing correction

Several of the autoimmune disorder groups that were significantly associated with AD risk in the UCSF data set were validated by the Stanford data set in one or both study designs. Gastrointestinal (case-control OR = 2.5, 95% CI 1.5-4.2, corrected p = 1.6e-3; cohort OR = 2.0, 95% CI 1.4-3.0, corrected p = 1.1e-3, Fig 3B), endocrine (cohort OR = 1.9, 95% CI 1.3-2.6, corrected p = 1.4e-3, Fig 3B), musculoskeletal (case-control OR = 1.7, 95% CI 1.3-2.3, corrected p = 1.6e-3; cohort OR = 1.5, 95% CI 1.2-1.8, corrected p=1.0e-3, Fig 3B), and dermatologic (case-control OR = 2.9, 95% CI 1.8-4.7, corrected p=1.7e-5; cohort OR = 1.6, 95% CI 1.2-2.2, corrected p = 2.6e-2, Fig 3B) disorders all conferred significantly more AD risk in autoimmune patients compared to non-autoimmune controls in the Stanford EHR system.

Next, we investigated if specific autoimmune disorders were driving the larger subtype and overall risk associations to determine if particular disorders were greater risk factors than others. Within the gastrointestinal category, inflammatory bowel disease was consistently associated with AD risk across UCSF (case-control OR = 5.0, 95% CI 2.1-12.9, corrected p=4.7e-4; cohort OR = 2.1, 95% CI 1.1-4.0, corrected p=0.050, Fig 3B, S3) and Stanford (case-control OR = 4.0, 95% CI 2.1-7.9, corrected p = 3.7e-5; cohort OR = 2.3, 95% CI 1.4-3.7, corrected p = 1.1e-3, Fig 3B, S3) study groups. Other disorders that conferred increased AD risk in at least two study groups across data sets included autoimmune thyroiditis, type 1 diabetes, and rheumatoid arthritis. Autoimmune thyroiditis, in the endocrine category, exhibited an odds ratio of 3.0 (95% CI 1.4-6.4, corrected p = 9.0e-3, Fig 3B, S3) in the case-control study group and 2.3 (95% CI 1.4-4.1, corrected p = 4.2e-3, Fig 3B, S3) in the cohort study group at UCSF, in addition to being a risk factor in the cohort study group of the Stanford data set (OR = 2.4, 95% CI 1.3-4.7, corrected p = 1.1e-2, Fig 3B, S3). Also in the endocrine category, type 1 diabetes was significantly associated with increased AD risk in the UCSF case-control study group (OR = 2.8, 95% CI 1.6-4.9, corrected p=8.6e-4, Fig 3B, S3) while also associating with nominally significant risk in the UCSF cohort study group (OR = 1.6, 95% CI 1.1-2.4, *uncorrected* p=0.02, Fig 3B, S3). Furthermore, type 1 diabetes was a significant AD risk factor in the Stanford cohort study group (OR = 1.8, 95% CI 1.2-2.7, corrected p=2.4e-2, Fig 3B, S3).

Validated drivers of risk like autoimmune thyroiditis and type 1 diabetes highlight a potential link between immune-mediated endocrine dysfunction and later AD pathogenesis. In addition to endocrine disorders, rheumatoid arthritis was the primary disease that conferred risk in the musculoskeletal disease category, and it was a significant risk factor in both UCSF study groups (case-control OR = 2.2, 95% CI 1.3-3.7, corrected p=1.1e-2; cohort OR = 2.1, 95% CI 1.4-3.2, corrected p=8.8e-4, Fig 3B, S3) as well as in the Stanford cohort study group (OR = 1.4, 95% CI 1.1-1.9, corrected p = 3.9e-2, Fig 3B, S3), with nominal significance in the Stanford case-control study group (OR = 1.5, 95% CI 1.0-2.3, *uncorrected* p = 2.9e-2, Fig 3B, S3).

### AD risk from disease subtypes exhibits variable sex-specific effects

Next, we stratified the disease subtype and individual disease analyses by sex to determine if any particular risk association was driven more by one sex compared to the other. Some disease subtypes were fairly constant in exhibiting what appeared to be sex-specific risk signals. For example, the endocrine category of disorders was predominantly significant among women, exhibiting a significant AD risk effect in the female UCSF study groups (case-control OR = 2.7, 95% CI 1.6-4.7, corrected p = 8.4e-4; cohort OR = 1.8, 95% CI 1.3-2.7, corrected p = 8.7e-3, Fig S2) and the female Stanford cohort study group (cohort OR = 1.8, 95% CI 1.2-2.8, corrected p = 0.04). In the dermatologic category, male-specific AD risk was higher than that of the female-specific comparison in the majority of study groups. In the UCSF cohort study group, men with dermatologic autoimmune disorders exhibited an odds ratio of 1.5 (95% CI 1.4-9.3, corrected p=0.04, Fig S2), while women exhibited an odds ratio of 2.0 that was insignificant after multiple testing correction (95% CI 1.1-3.8, corrected p=0.20, Fig S2). Similarly, in both Stanford study groups, men with dermatologic autoimmune disorders were at greater AD risk than controls (case-control OR = 7.8, 95% CI 3.4-18.5, corrected p=4.3e-7; cohort OR = 2.6, 95% CI 1.5-4.7, corrected p=1.9e-3, Fig S2) compared to women (case-control OR = 1.6, 95% CI 0.9-3.0, corrected p = 0.94; cohort OR = 1.2, 95% CI 0.8-1.8, corrected p = 3.5, Fig S2). Interestingly, in the case-control study group at UCSF, we also observed increased female-specific AD risk conferred by dermatologic autoimmune disorders (OR = 6.4, 95% CI 2.4-18.3, corrected p=4.4e-4, Fig S2), perhaps indicating variable sex-specific effects for this category of diseases. Similarly, we observed strong female-specific AD risk conferred by gastrointestinal conditions in our discovery data set (case-control OR = 11.8, 95% CI 4.2-37.1, corrected p=1.0e-6; cohort OR = 2.9, 95% CI 1.5-6.0, corrected p=6.5e-3, Fig S2), only to detect strong male-specific AD risk from these conditions in our validation data set (case-control OR = 3.4, 95% CI 1.4-8.5, corrected p=4.0e-2; cohort OR = 3.3, 95% CI 1.6-7.5, corrected p=4.8e-3, Fig S2). These findings warrant more investigation into the factors influencing the direction of sex-specificity for these disorder classes.

Clear sex-specific differences in the risk conferred by individual autoimmune disorders were harder to elucidate given a lack of power after stratifying by both sex and disorder (Table S2). We nonetheless were able to identify significant female-specific AD risk conferred by autoimmune thyroiditis in the cohort study groups of our two data sets (UCSF cohort OR = 2.4, 95% CI 1.3-4.5, corrected p=1.5e-2; Stanford cohort OR = 2.5, 95% CI 1.2-5.4, corrected p=3.1e-2, Fig S4), likely driving the female-specific risk in the endocrine category of diseases. Several other diseases increased AD risk significantly across some study groups, but not across others (Fig S4) or exhibited different directions of sex-specific effects across study groups (Fig S4). This again highlights the need for more studies of sex differences in the interaction between the autoimmunity and AD in the future.

### Sensitivity Analyses

We performed several sensitivity analyses to address the potential presence of confounders in EHR data and test the robustness of the risk associations we identified. The sensitivity analyses were performed in the discovery data set alone, as we wanted to verify the risk associations we saw in the smaller UCSF study groups before performing further validations in the Stanford study groups. Our first sensitivity analysis was to implement an age cutoff in our study designs to quantify AD risk due to autoimmunity in two older sub-populations of individuals. The first cutoff only included individuals in the study groups that had reached an age of 65 years or older. When comparing autoimmune patients to non-autoimmune controls in this >65 data set, we observed an AD odds ratio of 1.6 (95% CI 1.4-2.0, p=9.7e-9) in the case-control study group and 2.0 (95% CI 1.6-2.4, p=1.7e-14) in the cohort study group (Table S3). Raising the cutoff to 80 years of age or older also resulted in a strong AD risk association with autoimmune disorders (case-control OR = 1.7, 95% CI 1.4-2.2, p=1.7e-6; cohort OR = 2.4, 95% CI 1.8-3.1, p=1.8e-12, Table S3), indicating that our signal was robust to the age range of patients in our discovery data set.

Next we performed a sensitivity analysis to address possible confounder and collider effects that healthcare utilization can cause in EHR systems, since these effects can exaggerate or attenuate differences between exposure and outcome groups. In addition to matching on demographic variables (see Methods), we matched individuals based on the similarity of the length of time between their first and last hospital visit date. After conducting this matching and recomputing odds ratios in each study group, autoimmune disorders still associated with increased risk for AD (case-control OR = 1.3, 95% CI 1.1-1.5, p = 6.0e-3; cohort OR = 1.4, 95% CI 1.2-1.7, p = 5.2e-6, Table S3). Matching on each patient’s frequency of visits per year also resulted in consistent increased risk associations between autoimmune disorders and AD (case-control OR = 1.3, 95% CI 1.1-1.6, p = 3.7e-4; cohort OR = 1.5, 95% CI 1.3-1.7, p = 2.0e-6, Table S3), highlighting the connection between autoimmunity and AD at the clinical level even when adjusting for different healthcare utilization measures.

In the final sensitivity analysis, we recomputed odds ratios in each study group without including age at death as a variable in matching, to verify that leaving it out of the matching criteria did not significantly alter results (see Matching and Finalization of Study Groups in Methods). This resulted in an AD odds ratio of 1.3 (95% CI 1.1-1.5, p = 7.1e-4) in the case-control study group and 2.1 (95% CI 1.8-2.5, p = 4.7e-17) in the cohort study group (Table S3), indicating that leaving total lifespan information out of the matching criteria preserves risk signal. Several disease subtypes and individual disease associations seen in the main UCSF analysis were present in many, if not all, of these sensitivity conditions, including the increased AD risk due to endocrine, gastrointestinal, and musculoskeletal disorders driven by inflammatory bowel disease, type 1 diabetes, autoimmune thyroiditis, and rheumatoid arthritis (Table S3).

### Sex, but not autoimmunity, associates with accelerated AD onset over time

In addition to examining the presence or absence of AD in autoimmunity patients through our risk analyses, we also tested whether autoimmune disorders influence the timing of AD onset. We hypothesized that autoimmune disorders would decrease the age at which people are diagnosed with AD, potentially due to the early presence of chronic inflammation that autoimmune disorders bring about. We constructed new longitudinal cohorts for this analysis consisting only of AD patients with and without autoimmune disorders. This resulted in 292 autoimmune patients with 292 matched controls at UCSF (N = 584 total AD patients, Fig 4A), and 392 autoimmune patients with 392 matched controls at Stanford (N = 784 total AD patients, Fig 4A). We first compared the distributions of AD diagnosis age among patients. In the UCSF longitudinal cohort, the average age at which autoimmune patients were diagnosed with AD was 75.6 years, compared to the controls at 76.5 years (Fig S5A). In the Stanford longitudinal cohort, the average AD diagnosis age was 81.8 years in autoimmune patients compared to 82.4 years in controls (Fig S5A). While the AD diagnosis age was lower in autoimmune patients in each data set, the differences between age distributions were not significant in either case (UCSF p = 0.11, Stanford p = 0.17, Fig S5A), likely because of relatively small sample size (Table S2). The 0.6-to 1-year difference in diagnosis age we observed is nonetheless striking, given the often rapid symptomatic decline (21) of individuals with AD. Even being diagnosed with AD half a year earlier could be extremely impactful for patients and their quality of life.

**Figure 4.**
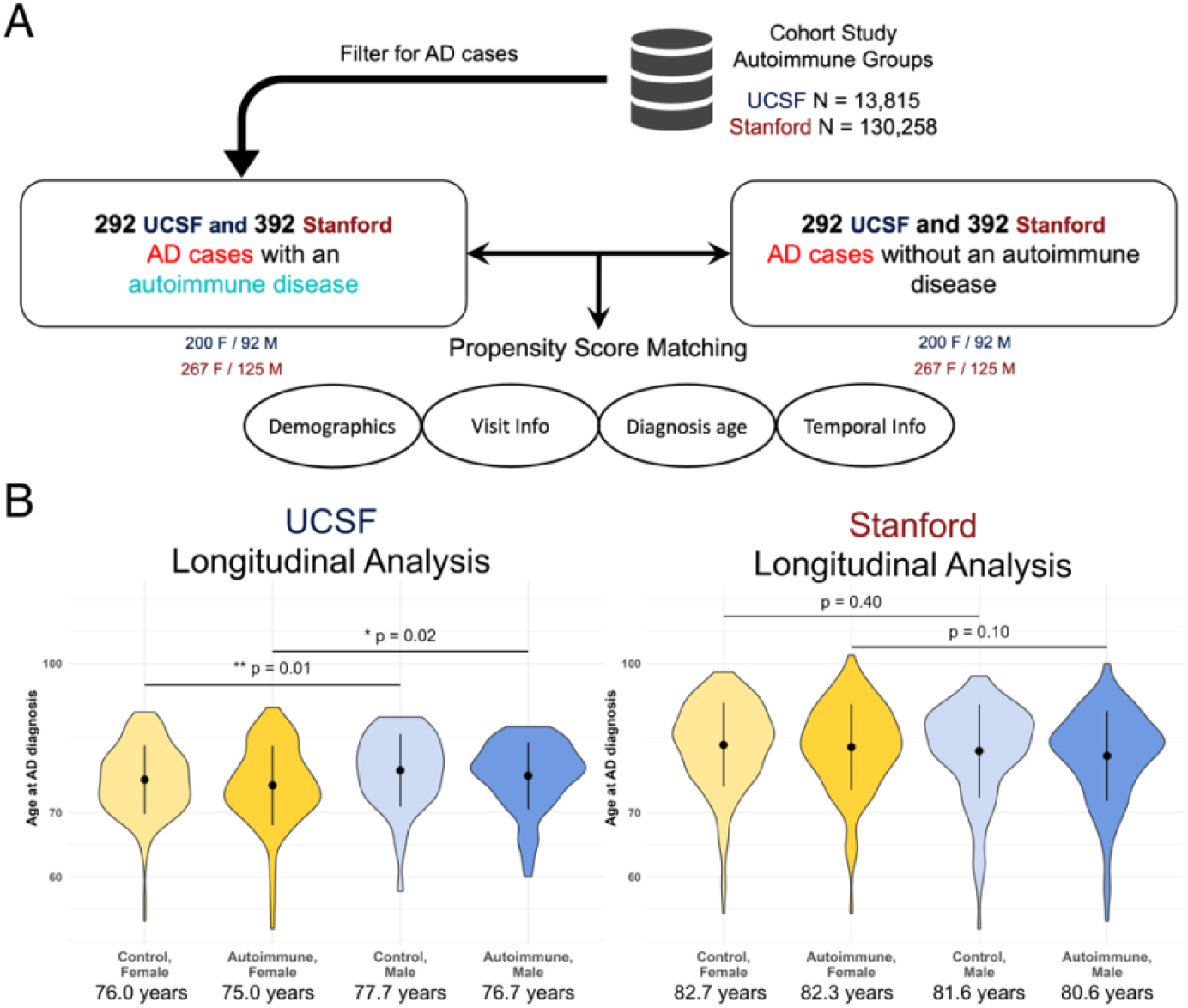
AD onset is accelerated in women, with potential acceleration from autoimmune disorders. **A)** Study design for the longitudinal AD onset analysis, conducted using data from the UCSF and Stanford EHR data sets. **B)** Distributions of AD diagnosis age among individuals with and without autoimmune diseases in the longitudinal cohorts, stratified by sex. Numbers below stratifications are the mean age at diagnosis, and the black summary lines highlight the mean and standard deviation of each distribution. Within the autoimmune and control subgroups, women were diagnosed with AD at a younger age than men, indicating that sex plays a large role in the age of AD onset.

We also observed differences in AD onset age when we stratified individuals both by sex and autoimmune disorder. Starting in the UCSF longitudinal cohort, women with autoimmune disorders exhibited significantly younger AD onset ages (mean 75.0 years, Fig 4B) compared to men with autoimmune disorders (mean 76.7 years, p = 0.017, Fig 4B). Similarly, control women exhibited significantly younger AD onset ages (mean 76.0 years, Fig 4B) compared to control men (mean 77.7 years, p = 0.011, Fig 4B). Comparing autoimmune patients to matched controls within each sex did not reveal younger onset ages (female p = 0.19, male p = 0.28, Fig 4B), likely indicating that sex plays a larger role in the timing of AD onset. Results from the Stanford longitudinal cohort all agreed directionally with the UCSF results, however with insignificant results (Fig 4B).

Finally, we performed a survival analysis to determine if the risk of developing AD in different stratifications varied over time. We used a Cox proportional hazard model for the analysis with sex and autoimmunity presence as covariates. Having an autoimmune disorder did not significantly alter the hazard of individuals developing AD over time (UCSF Hazard Ratio (HR) = 1.1, 95% CI 1.0-1.3, p = 0.16; Stanford HR = 1.0, 95% CI 0.9-1.2, p = 0.5, Fig S5B), corroborating the results from the distributional analysis. The hazard effects of sex were unclear given disagreement between UCSF and Stanford data sets. Being male in the UCSF longitudinal cohort trended toward being protective (HR = 0.85, 95% CI 0.7-1.0, p = 0.064, Fig S5B), while in the Stanford longitudinal cohort it was significantly associated with risk of developing AD earlier (HR = 1.2, 95% CI 1.0-1.4, p = 0.02, Fig S5B). Better powered analyses into sex as an influential variable in the timing of AD onset are needed to resolve this mismatch.

## Discussion

We have shown that autoimmune disorders are associated with increased risk of being diagnosed with AD in case-control and cohort study designs across two different EHR systems, suggesting that autoimmunity is a significant risk factor for AD. We observed a clear and strong increased risk signal in our study groups, both overall and in female- and male-specific subsets of our data, indicating that autoimmunity increases AD risk regardless of sex. Interestingly, in our AD prevalence analysis, women with autoimmune disorders exhibited increased AD prevalence compared to men with autoimmune disorders, indicating that both autoimmunity and sex play crucial roles in AD risk.

We observed specific subtypes of autoimmune disorders that conferred AD risk in patients. These particular classes of disorders suggest the presence of shared pathophysiology between particular autoimmunity subtypes and AD. For example, metabolic dysfunction has been highlighted in previous AD pathogenesis studies (22), so it is possible that metabolic pathways exist that link endocrine autoimmune disorders, which greatly affect metabolism, and AD pathogenesis. Similarly, there have been documented links between changes in the gut-brain axis (23) and microbiome (24) that are associated with AD in previous work, which could relate AD pathogenesis to pathways involved in gastrointestinal autoimmune disorders. Depending on the autoimmune disorder subtype and individual autoimmune disorders we analyzed, we also observed some sex-specific risk associations, highlighting that autoimmune disorders that fall under the category of certain physiological systems may manifest differently in women compared to men. This can provide more insight into the specific mechanisms that may go awry to cause or exacerbate AD in particular sexes.

Our findings validate and provide new insights on previous work. Recently, Miller et al. (25) found increased prevalence of inflammatory bowel disease in a small cohort of late-onset AD cases which we have validated in much larger clinical data sets. Furthermore, a study of the Swedish National Patient Register (26) highlighted increased incidence rates of several autoimmune disorders including hypothyroidism/thyroiditis, type 1 diabetes, Addison’s disease, Sjӧgren’s syndrome, and pernicious anemia in dementia patients. We were able to corroborate several of these signals with our analysis; we also found type 1 diabetes and a form of thyroiditis as significant modifiers of risk. We further built upon these findings by analyzing disease subtypes to identify particular physiological systems that may be more involved in AD pathogenesis, and we conducted extensive age of onset and sex-specific analyses to robustly characterize AD risk in autoimmune individuals. Finally, a recent study (27) in individuals of the UK Biobank indicated increased hazard ratios for four major autoimmune disorders (rheumatoid arthritis, multiple sclerosis, psoriasis, and inflammatory bowel disease) in a longitudinal cohort. Our results expand on this study by investigating several more autoimmune disorders and sex as a biological variable in the risk analyses. Compared to the individuals in the UK Biobank and Swedish studies, our population of patients was also more diverse, suggesting generalizability of the increased risk association between autoimmune disorders and AD across diverse populations of humans.

By using two different study designs (case-control and cohort) in both a discovery and validation data set in all of our analyses, we were further able to show robust associations while combating common problems of selection bias (28), data inaccuracies (29), and confounding that can be common in EHR studies. Our workflow can be further applied not only to autoimmune disorders and their risk relationship to other neurological diseases, but to any two disease types of interest that can be captured in an EHR data system.

The current study has several limitations that should be considered when evaluating our results. First, the groupings used for the disease subtype analysis are imperfect. Since the exact underlying mechanism of many of the autoimmune disorders we investigated remains unknown, grouping by physiological system enabled us to study these diseases in aggregate, but it reflects only one dimension along which each autoimmune disorder is related to the others. Another caveat is the presence of censored death information in the validation data set. This may have resulted in the incorporation of younger patients in the Stanford study groups who had yet to develop either AD or a later-onset autoimmune disorder, and as such, this may have deflated the odds ratios in the Stanford study groups. It is possible that this led to less agreement in risk associations between UCSF and Stanford particularly for the subtype and individual disorder analyses. While we matched individuals based on lifespan and/or birth year within each of our study groups, future work could include age-matching between discovery and validation data sets to enhance similarity of comparisons even further. Finally, the stratified analyses we conducted were less likely to yield consistent significant results across sites and study designs. Much of this was likely the result of reduced power due to the small sample sizes in the stratified cohorts. This also occasionally yielded very high odds ratios and large confidence intervals in our analyses. Nonetheless, many of these analyses highlight promising specific hypotheses for further validation and molecular characterization.

In summary, autoimmune disorders are strong risk factors for AD that act across sexes. This study illustrates the usefulness of EHRs for cross-trait analyses, and it also informs further mechanistic hypotheses about the exact molecular interfaces that may go awry in the interaction of the immune and nervous systems to promote AD pathogenesis. Further risk factor analyses for debilitating neurological conditions such as AD will empower clinicians to inform patients of their risk profiles for different diseases. Ultimately, deeper understanding of these connections between risk and disease can empower patients themselves to make lifestyle changes or take relevant treatments that can help avoid or delay disease. Our results highlight several future directions for further understanding of the risk relationship between autoimmune disorders and AD. First, quantifying how AD risk varies based on differing levels of autoimmune disorder severity and duration is needed. We hypothesize that more severe forms of autoimmune disorders may confer the most AD risk, and perhaps treatments to alleviate more severe disorders may decrease risk. Additionally, taking into account the chronic nature and onset age of many autoimmune disorders may shed more light on the temporal dynamics of the two traits interacting throughout the lifespan. Finally, integrating clinical risk analyses with other data modalities, such as genetics or proteomics will provide more molecular insight into the link between AD and autoimmune disorders and help to fully elucidate the basis for the strong risk seen at the phenotypic level in human populations.

## Methods

### Sex as a Biological Variable

We accounted for sex as a biological variable by performing all analyses in female- and male-specific subsets of our original study groups. This allowed us to test if any risk associations differed by sex, and if particular autoimmunity subtypes or individual disorders were driving AD risk in a particular sex. We filtered out individuals with an unknown sex from our UCSF study groups. Due to differences in encoding sex within the UCSF and Stanford EHR systems, there were a small number of individuals (0.02% of the total) with an unknown sex that were included in the Stanford cohort data set. These individuals were not included in any sex-specific analyses.

### Study Group Identification

#### Autoimmune Disorder, Alzheimer’s Disease, and Healthy Control Patient Identification

We identified 26 autoimmune disorders of interest (Table S1) for our study based on prior literature and prevalence in the general population. Individuals with each autoimmune disorder were identified by string-matching autoimmune disorder names (Table S4) with billing concepts. Concepts were subsequently standardized for use in the UCSF EHR database (30) which is based on the Observational Medical Outcomes Partnership (OMOP) Common Data Model (CDM) and primarily uses Systemized Nomenclature of Medicine (SNOMED) concept encodings. All standardized billing concepts were examined by UCSF rheumatologists to confirm validity and relevance of each concept to each autoimmune disorder of interest. We compiled a final list of 878 autoimmune billing concepts (Table S5). We identified patients who had these concepts present in their UCSF medical record to construct our discovery data set, and used identical concepts to identify patients from the Stanford EHR database for our validation data set. We identified patients with AD in a similar manner to autoimmune disorder patients after string-matching AD terms to concepts (Table S6) and checking for billing concept occurrence in each patient record in the UCSF and Stanford systems. We only included AD billing concepts related to late-onset or sporadic AD, as early-onset AD is thought to have distinct etiology and stronger genetic components (31). Additional demographic data from the UCSF and Stanford EHR systems was collected on patients including date of birth, date of death (if available), self-reported race, self-reported ethnicity, and sex. We also gathered healthcare utilization statistics on patients, including number of doctor’s visits, total number of unique diagnoses, and first and last medical visit date.

We identified healthy control individuals without autoimmune disorders to compare to the autoimmune patient groups from the UCSF and Stanford databases. To do this, we searched for patients *without* any of the 878 finalized autoimmune disorder billing concepts present in their records, and we additionally removed individuals from the healthy control group who had concepts that were similar to any of the 878 finalized autoimmune concepts. For example, an individual who had the billing concept “family history of Celiac disease” in their EHR but did not have a more specific Celiac disease billing concept identifying a personal diagnosis of Celiac disease would have been excluded. Similarly, several billing concepts that were too general to pertain specifically to an autoimmune disorder (e.g. kidney disease) but that represented a serious condition were excluded from the healthy controls. For our cohort study design, we matched the healthy controls to autoimmune disorder patients based on criteria further described in the *Matching and Finalization of Study Groups* section of the Methods.

To identify a population of non-AD healthy controls to compare to our AD patient group, we searched for patients *without* any AD billing concepts in their records and matched them with AD cases based on demographic factors. To clarify some terminology, the control individuals that were matched to autoimmune disorder cases will be referred to as the “non-autoimmune controls” going forward, whereas the control individuals that were matched to AD cases will be referred to as the “non-AD controls”. These are two separate groups of controls, but they may have overlap in individuals as someone without both an autoimmune disorder and AD might be in both control groups. Additionally, someone in one of the disease groups may be in a control group for the other disease. For example, an AD patient might show up in the non-autoimmune control group, since the requirement to be in that group is not having an autoimmune disorder.

#### Data Cleaning and Quality Control

Our data cleaning pipeline involved several steps. First, we performed quality control to remove any individuals with missing demographic information in the self-reported race, self-reported ethnicity, sex, and birth year categories from consideration for our disease and healthy control groups in the UCSF data set. Due to differences in encoding some of this demographic information between UCSF and Stanford EHR systems, a small number of individuals with unknown demographic values were included in the Stanford data set (see Sex as a Biological Variable), but each individual with an “unknown” demographic field was similarly matched with another Stanford individual with an “unknown” value, removing any issues comparing people without matching information. In the UCSF study groups, we also required individuals to have a valid reported age at death, as this allowed us to compare individuals by total lifespan. We restricted individuals to be between 30 and 90 years of age at their death in these groups. In the Stanford study groups, there was a greater degree of censored death information such that including lifespan information in matching would have extremely limited our study group sizes, so we did not enforce this constraint. We verified that leaving out lifespan as a matching variable in the Stanford groups did not significantly alter overall risk signals, so we felt confident leaving it out of our validation study group criteria (See Sensitivity Analyses in Results). It is likely that, because of censored data being included in the Stanford data sets, the increased risk associations we saw in our analyses would be even stronger in real life (see Discussion). In addition to quality control on demographics, we also removed individuals who had zero hospital visits or whose first and last visit dates were the same.

We next determined 1) who in the autoimmune disorder patient groups and corresponding non-autoimmune control groups had an AD diagnosis, and 2) who in the AD patient groups and corresponding non-AD control groups had an autoimmune disorder diagnosis. Within our autoimmune/AD disease groups and respective healthy control groups, different individuals were demarcated with their assigned “person ID” following OMOP conventions. We then determined which person IDs of one group overlapped with the person IDs of another group. For example, to discover which autoimmune patients and non-autoimmune controls had an AD diagnosis, we determined which of the person IDs of our AD patient group overlapped with the person IDs of the autoimmune and non-autoimmune groups. In a similar manner, we determined which AD patients and which non-AD controls had an autoimmune diagnosis by overlapping the person IDs of our autoimmune patient group with the AD and non-AD person IDs.

We also identified the relative dates of AD and autoimmunity diagnoses for patients. To focus on the effect of autoimmunity on AD, we did not consider individuals who had an AD diagnosis prior to their autoimmune disorder diagnosis. Specifically in the UCSF data set, we computed several metrics for each individual to aid the matching of autoimmune or AD patients to their respective controls when performing different sensitivity analyses. These metrics included each individual’s age when they died, the total length of the UCSF EHR record (last visit date -first visit date), and the frequency of doctor’s visits per year

To understand which autoimmune disorder subtypes might be driving AD risk, we grouped the 26 autoimmune disorders of interest into 8 distinct subtype categories (Table S1 and Fig 3A) and assigned patients into categories based on which autoimmune disorder(s) they had. If a patient had multiple autoimmune disorders (UCSF N = 2,378 patients, Stanford N = 37,332 patients) across different subtype categories, they were counted once within each subtype risk association analysis, and therefore could be represented in more than one analysis.

#### Study Designs, Matching, and Finalization of Study Groups

To mitigate selection bias and ensure robustness of results, we examined the risk of receiving an AD diagnosis following an autoimmune disorder diagnosis using two study designs in each of our EHR data sets: a retrospective case-control study with AD patients and non-AD matched controls, and a retrospective cohort study with autoimmune disorder patients and non-autoimmune matched controls. We performed 1:1 matching of patients to controls for each study group using propensity scoring on each individual’s birth year, sex, self-reported race, and self-reported ethnicity. We also matched on lifespan in the UCSF study groups (See Data Cleaning). We enforced exact matches between patients and controls in the categories of sex, self-reported race, and self-reported ethnicity. In our main analysis and throughout follow-up sensitivity analyses, we ensured high-quality matching by verifying that the average absolute standardized mean error between each matched pair was less than 0.1. We conducted final quality control in both study groups of our main and sensitivity analyses by removing any matched pairs of individuals where a control individual was diagnosed with AD prior to the matched disease case being diagnosed with an autoimmune disorder. All matching was performed using the nearest neighbor method of the MatchIt package (4.5.3) in R. After matching and cleaning, we were left with four “study groups”: a case-control and cohort study group from UCSF, and a case-control and cohort study group from Stanford.

### AD Risk Analysis in Case-Control and Cohort Study Groups

In order to enable comparison across the different study designs, we computed odds ratios to quantify the risk of being diagnosed with AD in autoimmune disorder patients compared to non-autoimmune controls. Across each of the study groups, we computed odds ratios at three levels: 1) across all autoimmune disorders combined, 2) across autoimmunity subtypes, and 3) across individual autoimmune disorders. At each of these three levels, we repeated the analysis in a sex-stratified manner to explore if risk was sex-specific. All odds ratios were computed using a Fisher exact test on contingency tables of AD/autoimmunity patients, and we used the oddsratio.fisher function of the epitools package (version 0.5-10.1) in R to compute these statistics.

To account for multiple hypothesis testing, we used Bonferroni corrections for each odds ratio analysis. For the disease subtype risk analysis, we corrected p-values within each overall or sex stratification group (e.g., a correction factor of 8 for the 8 disease subtype comparisons). For the specific disorder risk analysis, we performed a within-disorder-subtype p-value correction, to evaluate which specific conditions within an autoimmune disorder subtype group had significant subtype effects. This meant the correction factor for a particular disorder comparison was determined by the number of conditions within that particular disorder’s subtype category (e.g., p-values for each autoimmune disorder in the endocrine subgroup were corrected by a factor of 4, due to the endocrine subgroup being comprised of 4 diseases).

### AD Prevalence Calculations in Cohort Study Groups

We calculated the prevalence of AD in patients with autoimmune disorders compared to matched controls in a sex-stratified manner. To ensure that a difference in prevalence was not due to underlying demographic differences between female and male individuals, we took the cohort study groups across UCSF and Stanford and matched the smaller sample size of male individuals to female individuals based on birth year, self-reported race, and self-reported ethnicity, while matching additionally on lifespan in the UCSF study groups. Again, we used exact matching on self-reported race and self-reported ethnicity while using propensity score matching on the remaining variables. We then calculated the percentage of people with AD in each stratification: female individuals with autoimmune disorders, male individuals with autoimmune disorders, female non-autoimmune control individuals, and male non-autoimmune control individuals. To obtain a 95% confidence interval for each prevalence statistic, we bootstrapped the data 1,000 times. We again used a Fisher’s exact test to compute significance of prevalence differences among stratifications.

### Longitudinal AD Onset Analysis

To understand the effect of autoimmunity on the risk of AD diagnosis over time, we conducted a longitudinal age of onset analysis. For this, we constructed new longitudinal study groups from the UCSF and Stanford EHR systems consisting of individuals with both an autoimmune disorder and AD from our cohort study designs. These longitudinal cohorts also passed our data quality control pipeline. We matched the autoimmunity patients with AD to non-autoimmune individuals with AD from the larger UCSF and Stanford background databases. The same variables used in matching for the main odds ratio analysis were also used here. The age of AD onset for each individual in each longitudinal study group was then calculated by taking the difference in time between a person’s birth year and the first appearance of an AD billing concept in the person’s medical record. The Mann–Whitney U test and Cox proportional hazard modeling conducted in these study groups was performed using the stats (v4.1.3) and survival/survminer (v3.5-5/0.4.9) packages, respectively, in R.

### Statistics

We used Fisher’s exact test for all odds ratio calculations and for comparisons in our AD prevalence analysis. We also used a Mann-Whitney U test for a non-parametric comparison of distributions in our longitudinal AD onset analysis. Bonferroni corrections were applied to p values throughout this study; these are denoted by the use of “corrected” or “adjusted” p values.

### Study Approval

All analysis of University of California, San Francisco and Stanford University electronic health record data was performed under the approval of the Institutional Review Boards from University of California, San Francisco and Stanford University, respectively. All clinical data were de-identified and written informed consent was waived by the institutions.

## Supporting information

Supporting Data Values

## Data Availability

Individual patient data is not publicly available due to patient data sharing privacy. Code not limited by patient data sharing permissions can be found at https://github.com/gramey02/AD_AID_Project. All patient and demographic data curation from the UCSF and Stanford EHR systems was performed using Microsoft SQL server and the DBI (v1.1.3) and odbc (1.3.4) packages in R. Discovery data was last curated from the UCSF OMOP database on August 4th, 2023, and validation data was last curated from the Stanford OMOP database on December 12th, 2023. Data cleaning, matching, and analysis steps were conducted using R version 4.1.3, and plots were created with the ggplot2 package (v3.4.2). Values for all data points in graphs are reported in the Supporting Data Values file.

## Author Contributions

A.T., Z.M., M.S, and G.D.R. conceptualized this research project. G.D.R. investigated and curated all UCSF data and performed all UCSF statistical and computational analysis. T.P. performed all Stanford validation steps, including data curation and statistical/computational analysis. M.Y. provided valuable clinical input on EHR methodology and clinical billing codes. G.D.R. and A.T. wrote the manuscript, and G.D.R. created visualizations. J.A.C. and M.S. provided mentorship, review/editing of writing, and funding for the work. I.A., N.A., T.M., T.O., and S.W. provided expertise and manuscript feedback.

## Acknowledgements

We would like to thank all of the members of J.A.C.’s, M.S.’s, and N.A.’s labs for all of their feedback throughout this work. We acknowledge our funding sources, including NIA R01AG060393, NIAMS P30 AR070155, F30 Fellowship 1F30AG079504-01, and the UCSF Discovery Fellows. We would also like to acknowledge the use of the [UCSF Information Commons](https://informationcommons.ucsf.edu/) and [UCSF Research Analysis Environment](https://it.ucsf.edu/service/rae) computational research platforms. Through these platforms, the project was supported by the National Center for Advancing Translational Sciences, National Institutes of Health, through UCSF-CTSI Grant Number UL1 TR001872. Its contents are solely the responsibility of the authors and do not necessarily represent the official views of the NIH.

