## Supplementary Figures for "Exposure to autoimmune disorders increases Alzheimer’s disease risk in a multi-site electronic health record analysis"

### Supplemental Figures and Figure Legends

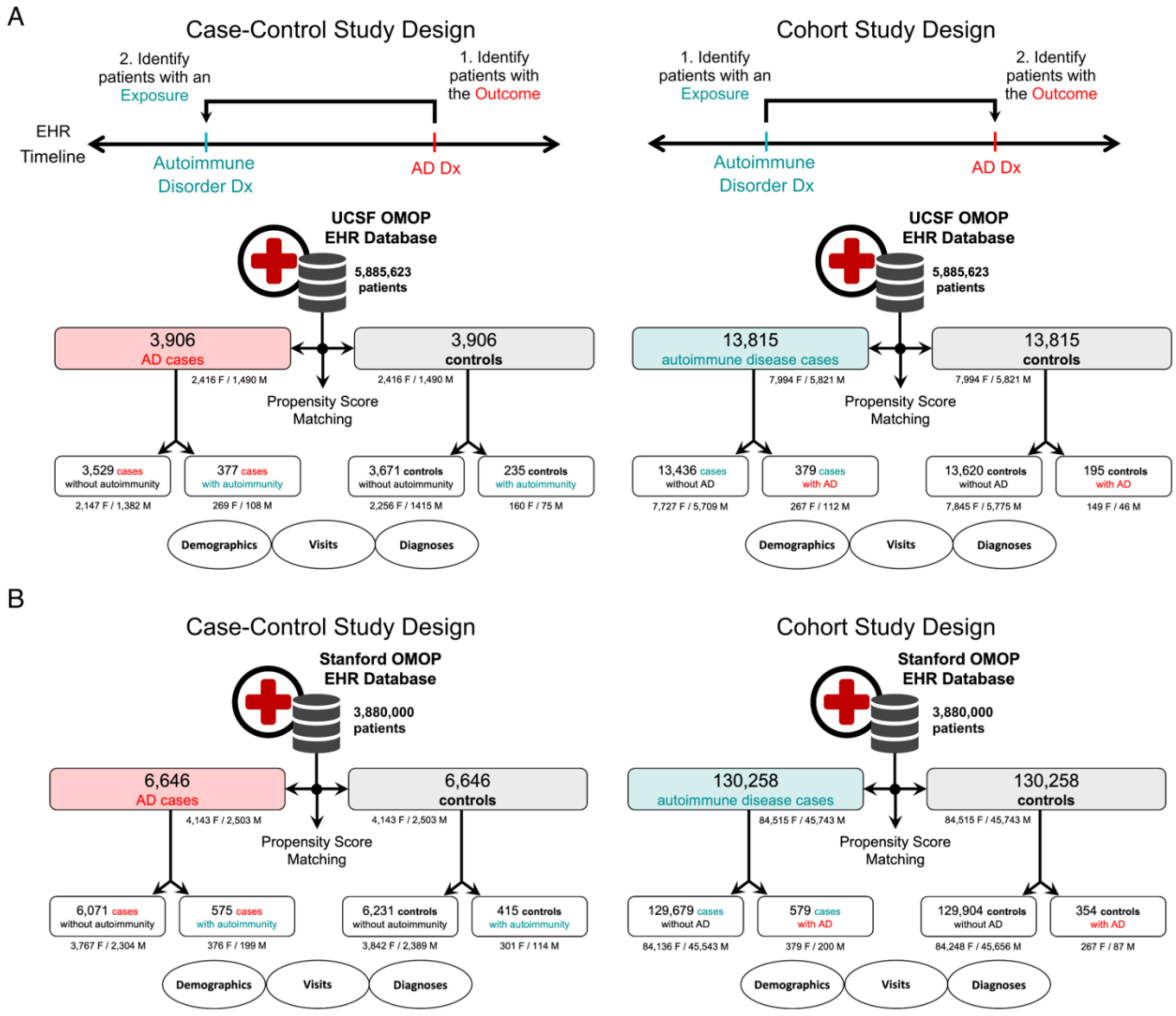


**Figure S1: Full breakdown of study groups in UCSF and Stanford data sets.** Numbers of patients with autoimmune disorders and/or AD in the case-control (left) and cohort (right) study groups of either the UCSF (A) or Stanford (B) data sets. Also pictured are the breakdowns of each category by sex. Dx = Diagnosis, M = Male, F = Female

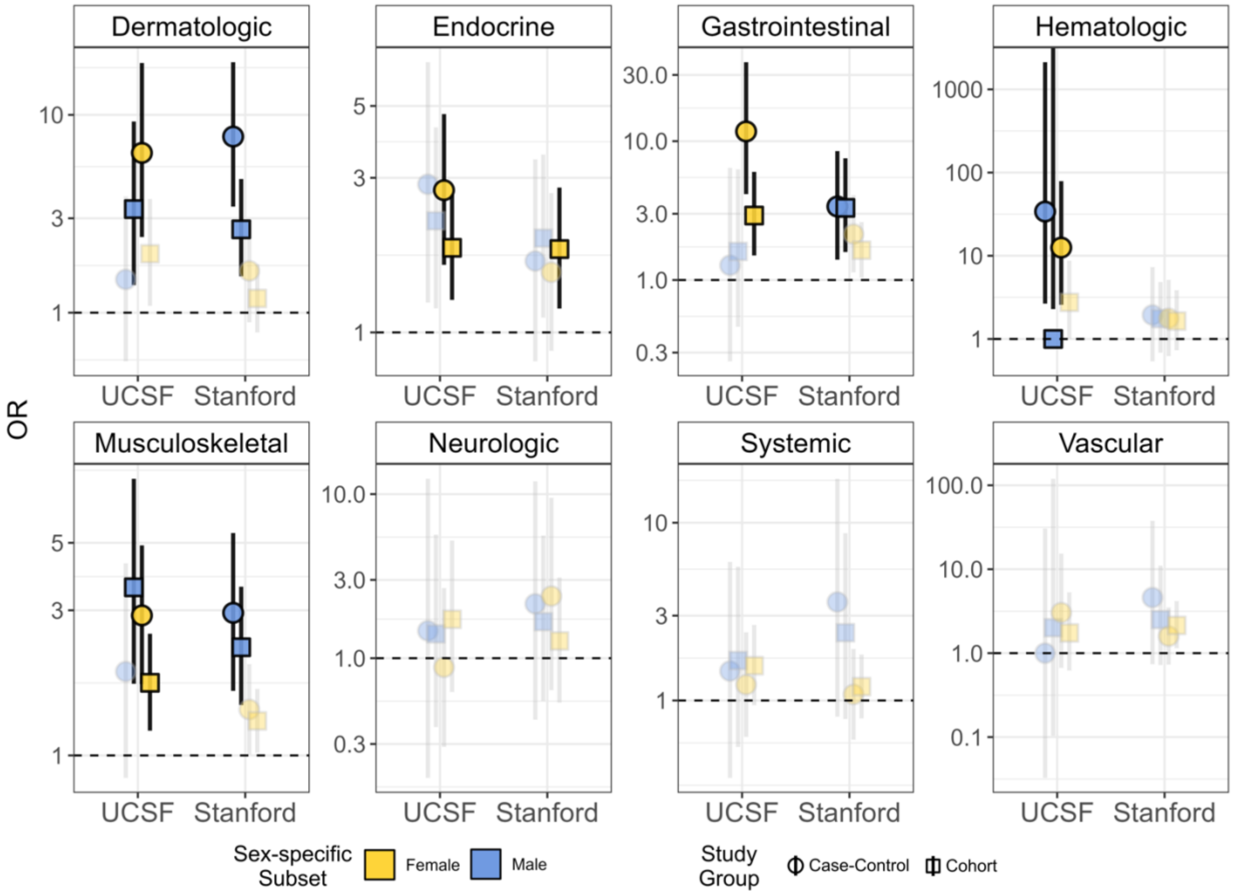


**Figure S2: Autoimmune disorder subtypes exhibit varying sex-specific AD risk effects.** AD odds ratios for each of the eight autoimmune disorder subtypes across female-and male-specific subsets of each EHR system and study group. Significant odds ratios are bold/outlined in black while insignificant odds ratios are faded/more transparent. Many subtypes associated with increased AD risk predominantly in one sex more than another (e.g. dermatologic disorders in males and endocrine disorders in females) whereas other subtypes were less clear in their sex-specific effects (e.g. gastrointestinal and musculoskeletal conditions), which varied between discovery and validation data sets.

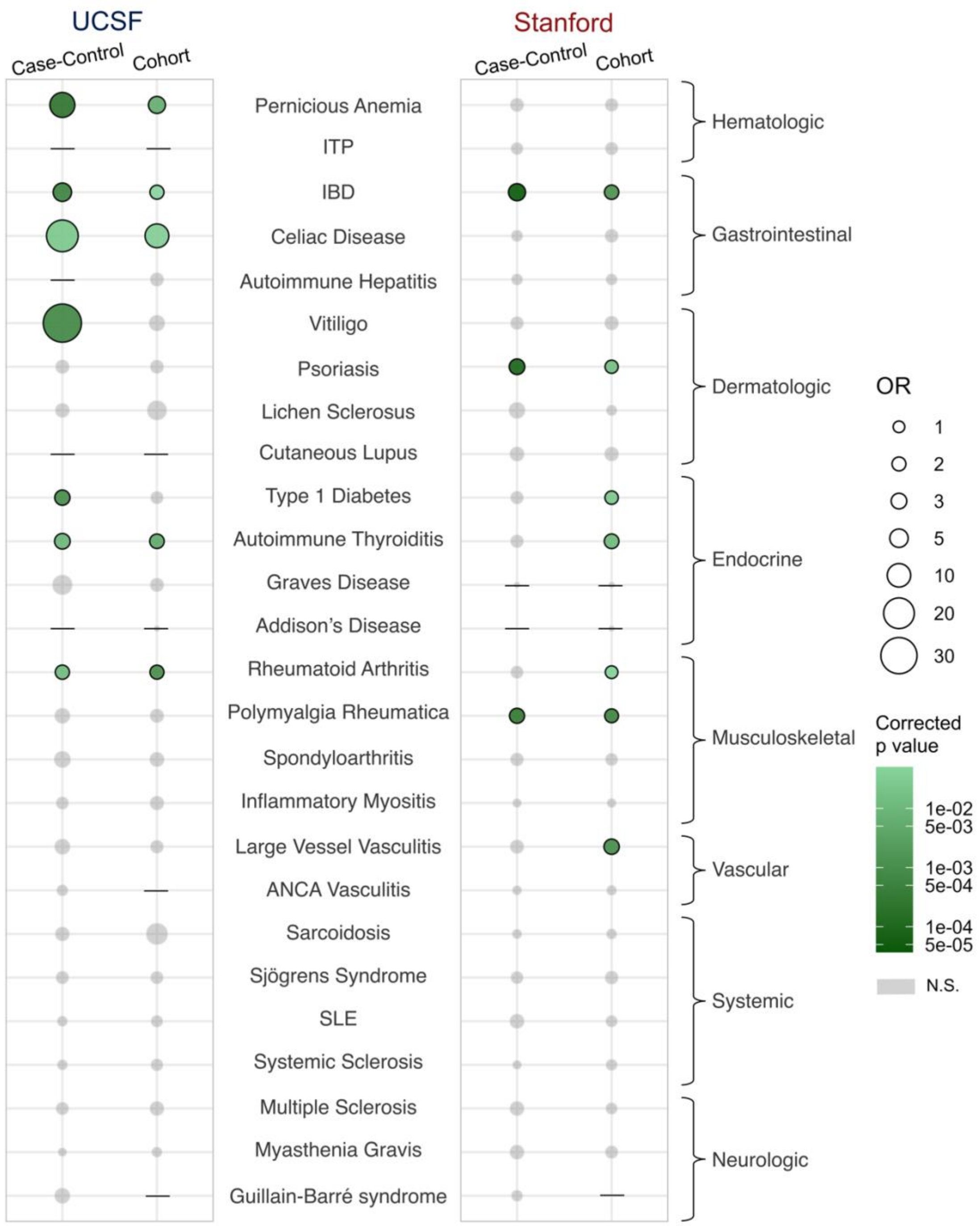


**Figure S3: Specific autoimmune disorders are associated with increased AD risk.** AD odds ratios for each of the 26 specific autoimmune disorders we analyzed. Many were significant across several study designs and data sets, including IBD, type 1 diabetes, autoimmune thyroiditis, and rheumatoid arthritis. For several of the more rare autoimmune disorders, we were underpowered to detect risk associations, and those with sample sizes that were too small for calculations (e.g. zero AD cases) or resulted in uninterpretable estimations (e.g. infinite odds ratio or confidence interval estimation) are denoted with black horizontal line segments. These diseases are nonetheless interesting and warrant more research. ITP = Idiopathic Thrombocytopenic Purpura, IBD = Inflammatory Bowel Disease, SLE = Systemic Lupus Erythematosus, N.S. = Not Significant

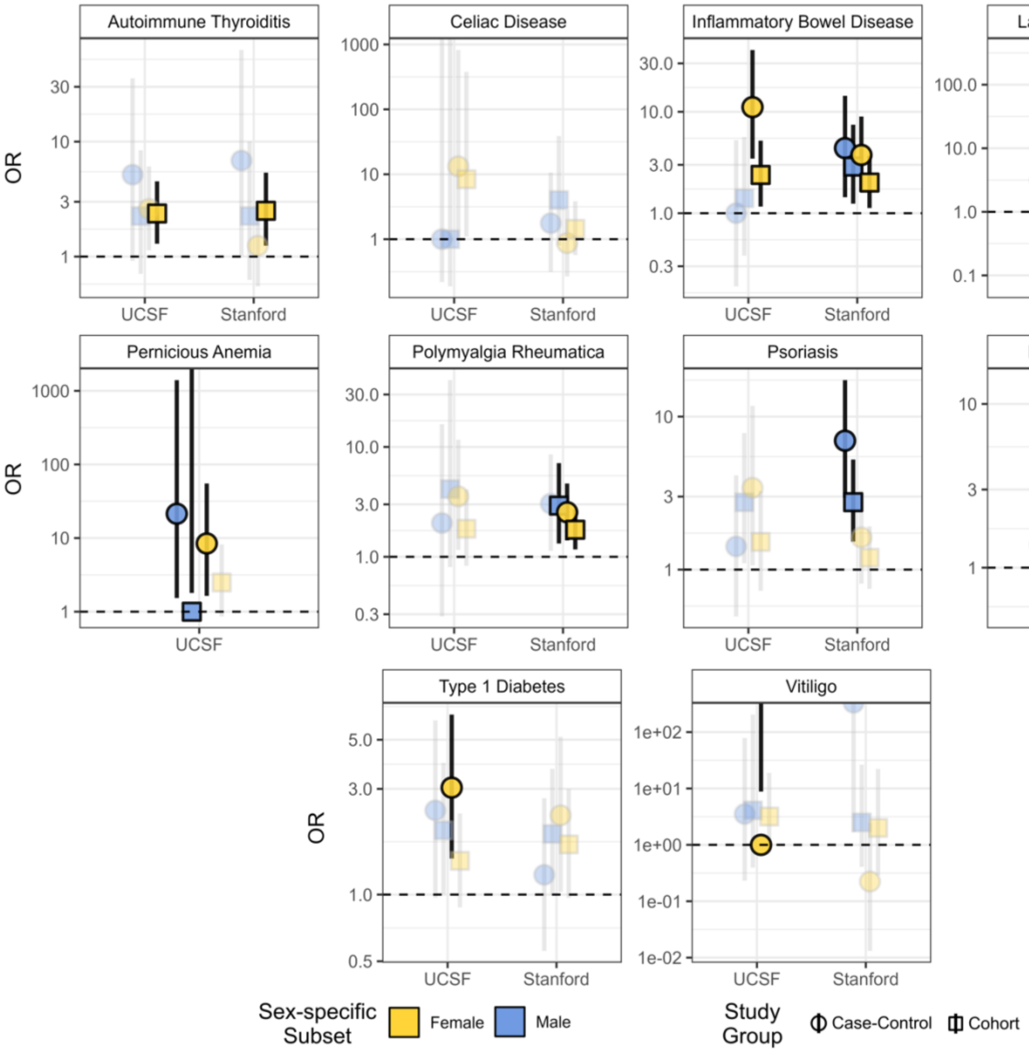


**Figure S4: Increased AD risk from specific autoimmune disorders is potentially sex-specific.** Sex-specific AD odds ratios for each of the specific autoimmune disorders that were significant across multiple data sets in the overall analysis (Fig 3B). Significant odds ratios are outlined in black while insignificant associations are faded/more transparent. Autoimmune thyroiditis increased risk primarily in women, potentially driving the female-specific AD risk association for endocrine diseases (Fig S2). Given that we were underpowered to detect several of these associations after stratifying at so many levels, more research into the sex-specific effects of particular autoimmune disorders is warranted, especially given the sex disparities that remain despite increased risk across both sexes overall. Odds ratios pictured that fall outside of their respective confidence intervals were biologically uninterpretable due to infinitely high confidence intervals and/or odds ratios.

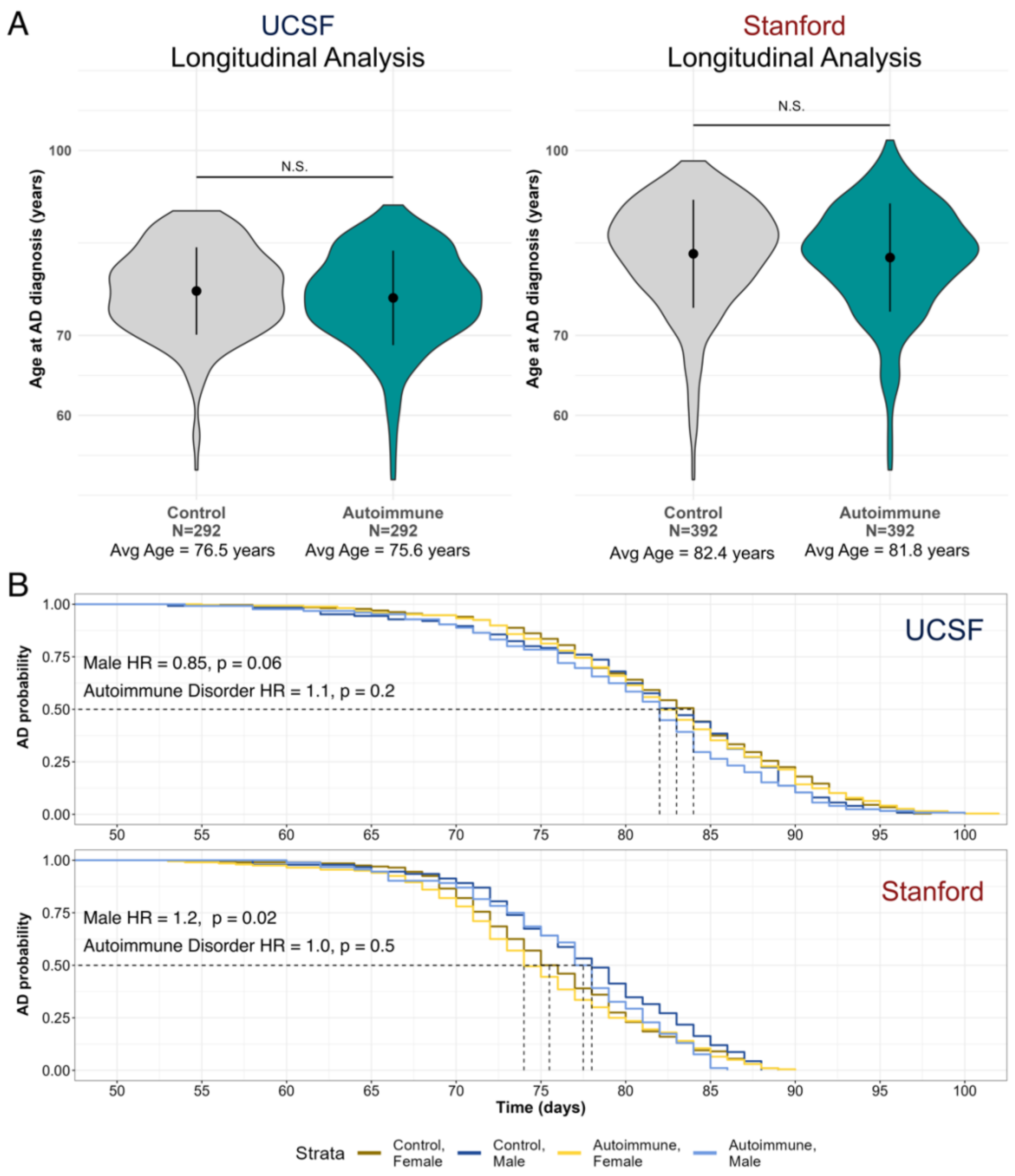


**Figure S5: Autoimmune disorders trend toward accelerating AD onset.** A) Distributions of AD diagnosis age among individuals with autoimmune disorders and non-autoimmune controls in UCSF (left) and Stanford (right) longitudinal cohorts. While younger age of onset in autoimmune patients was insignificant compared to controls, likely due to power (see B), there was about a one-year difference between autoimmunity cases and controls. B) Survival curves for individuals in the longitudinal study groups at UCSF (top) and Stanford (bottom). Like in the distributional analysis, sex tended to separate individuals into groups of relatively earlier or later AD onset, while the presence of an autoimmune disorder did not.
